# Automated diagnosis of atrial fibrillation in 24-hour Holter recording based on deep learning:a study with randomized and real-world data validation

**DOI:** 10.1101/2021.08.25.21262591

**Authors:** Peng Zhang, Fan Lin, Fei Ma, Yuting Chen, Daowen Wang, Xiaoyun Yang, Qiang Li

**Author notes:** Correspondence to: Qiang Li, PhD, Wuhan National Laboratory for Optoelectronics, Huazhong University of Science and Technology, 1037 Luoyu Road, Wuhan, Hubei, China, Xiaoyun Yang, PhD, Division of Cardiology, Department of Internal Medicine, Tongji Hospital, Tongji, Medical College, Huazhong University of Science and Technology, Wuhan, Hubei, China. Drs. Zhang and Lin contributed equally.

## Abstract

**Background:** With the increasing demand for atrial fibrillation (AF) screening, clinicians spend a significant amount of time in identifying the AF signals from massive electrocardiogram (ECG) data in long-term dynamic ECG monitoring. In this study, we aim to reduce clinicians’ workload and promote AF screening by using artificial intelligence (AI) to automatically detect AF episodes and identify AF patients in 24 h Holter recording.

**Methods:** We used a total of 22 979 Holter recordings (24 h) from 22 757 adult patients and established accurate annotations for AF by cardiologists. First, a randomized clinical cohort of 3 000 recordings (1 500 AF and 1 500 non-AF) from 3000 patients recorded between April 2012 and May 2020 was collected and randomly divided into training, validation and test sets (10:1:4). Then, a deep-learning-based AI model was developed to automatically detect AF episode using RR intervals and was tested with the test set. Based on AF episode detection results, AF patients were automatically identified by using a criterion of at least one AF episode of 6 min or longer. Finally, the clinical effectiveness of the model was verified with an independent real-world test set including 19 979 recordings (1 006 AF and 18 973 non-AF) from 19 757 consecutive patients recorded between June 2020 and January 2021.

**Findings:** Our model achieved high performance for AF episode detection in both test sets (sensitivity: 0.992 and 0.972; specificity: 0.997 and 0.997, respectively). It also achieved high performance for AF patient identification in both test sets (sensitivity:0.993 and 0.994; specificity: 0.990 and 0.973, respectively). Moreover, it obtained superior and consistent performance in an external public database.

**Interpretation:** Our AI model can automatically identify AF in long-term ECG recording with high accuracy. This cost-effective strategy may promote AF screening by improving diagnostic effectiveness and reducing clinical workload.

**Research in context:** *Evidence before this study:* We searched Google Scholar and PubMed for research articles on artificial intelligence-based diagnosis of atrial fibrillation (AF) published in English between Jan 1, 2016 and Aug 1, 2021, using the search terms “deep learning” OR “deep neural network” OR “machine learning” OR “artificial intelligence” AND “atrial fibrillation”. We found that most of the previous deep learning models in AF detection were trained and validated on benchmark datasets (such as the PhysioNet database, the Massachusetts Institute of Technology Beth Israel Hospital AF database or Long-Term AF database), in which there were less than 100 patients or the recordings contained only short ECG segments (30-60s). Our search did not identify any articles that explored deep neural networks for AF detection in large real-world dataset of 24 h Holter recording, nor did we find articles that can automatically identify patients with AF in 24 h Holter recording.

*Added value of this study:* First, long-term Holter monitoring is the main method of AF screening, however, most previous studies of automatic AF detection mainly tested on short ECG recordings. This work focused on 24 h Holter recording data and achieved high accuracy in detecting AF episodes. Second, AF episodes detection did not automatically transform to AF patient identification in 24 h Holter recording, since at present, there is no well-recognized criterion for automatically identifying AF patient. Therefore, we established a criterion to identify AF patients by use of at least one AF episode of 6 min or longer, as this condition led to significantly increased risk of thromboembolism. Using this criterion, our method identified AF patients with high accuracy. Finally, and more importantly, our model was trained on a randomized clinical dataset and tested on an independent real-world clinical dataset to show great potential in clinical application. We did not exclude rare or special cases in the real-world dataset so as not to inflate our AF detection performance. To the best of our knowledge, this is the first study to automatically identifies both AF episodes and AF patients in 24 h Holter recording of large real-world clinical dataset.

*Implications of all the available evidence:* Our deep learning model automatically identified AF patient with high accuracy in 24 h Holter recording and was verified in real-world data, therefore, it can be embedded into the Holter analysis system and deployed at the clinical level to assist the decision making of Holter analysis system and clinicians. This approach can help improve the efficiency of AF screening and reduce the cost for AF diagnosis. In addition, our RR-interval-based model achieved comparable or better performance than the raw-ECG-based method, and can be widely applied to medical devices that can collect heartbeat information, including not only the multi-lead and single-lead Holter devices, but also other wearable devices that can reliably measure the heartbeat signals.

## Introduction

Atrial fibrillation (AF) is the most common tachyarrhythmia, with a prevalence of 1-2% in the general population, and it can lead to dangerous complications and increase cardiovascular mortality risk^1^. AF can be classified into three types depending on the episode duration, namely, paroxysmal AF (PAF), persistent AF, and permanent AF^2^. PAF implies to occasional or intermittent episodes and is difficult to detect with resting ECG or short-term ECG monitoring. In clinical practice, long-term dynamic ECG monitoring (Holter monitoring) is the main method to detect PAF^3^. However, long-term Holter monitoring produces a large amount of ECG data and thus brings heavy workloads to clinicians in identifying the AF signals. Moreover, the identification of AF signals is subjective and heavily depends on the experience of clinicians. However, experienced cardiologists are scarce, especially in low- and middle-income countries^4^. Therefore, it is particularly important to develop an artificial intelligence (AI)-based AF diagnostic framework to reduce the subjective variabilities and the workload in identifying AF signals in Holter monitoring.

In recent years, many deep-learning-based methods have been proposed for automatic AF detection and achieved good performance in benchmark datasets^5-8^, however, three major problems remain. First, 24 h Holter recording is the main method for AF screening, but is rarely employed in automated diagnosis of AF. Most studies focused on short ECG recordings with duration of a few minutes, up to 10 h^9^. Second, the verification of automated AF detection in real-world environments is scarce^10^. Most studies used public datasets only, such as the PhysioNet 2017 database^11^, MIT-BIH AF database^12^, and Long-Term AF database^13^, which included only short ECG recordings or a small number of patients. Although a few studies evaluated deep-learning-based automated ECG analysis with large clinical datasets, these studies aimed to classify multiple types of arrhythmias and did not achieve high enough accuracy in detecting AF episode^14-16^. Finally, the automatic detection of AF episode in previous studies can not realize the automatic screening of AF in long-term Holter recording, and the combination with automatic identification of AF patient is further required.

To address these challenges, we developed an AI pipeline for automatic AF episode detection and AF patient identification in 24 h Holter recording, further evaluated its effectiveness using randomized and real-world clinical datasets, and discussed its applicability in clinical practice. To the best of our knowledge, this is the first study to use large 24 h Holter recording dataset collected from the real-world clinical practice to test an automated AF diagnosis model.

## Methods

### Study design

Our primary goal was to develop an AI pipeline for automatic (a) AF episode detection and (b) AF patient identification in Holter recording; the pipeline used deep learning technology and required no patient or clinician intervention. Our method contained multiple steps as shown in **Figure. 1**. From 24 h Holter recording, 12-lead ECG raw data were obtained and used to extract 90-RR-interval samples. Then the deep neural network detected each 90-RR-interval sample as AF or non-AF (NAF). Finally, based on the results of AF episode detection, AF patients were automatically identified using our proposed criterion of at least one AF episode of 6 min or longer.

**Figure. 1.**
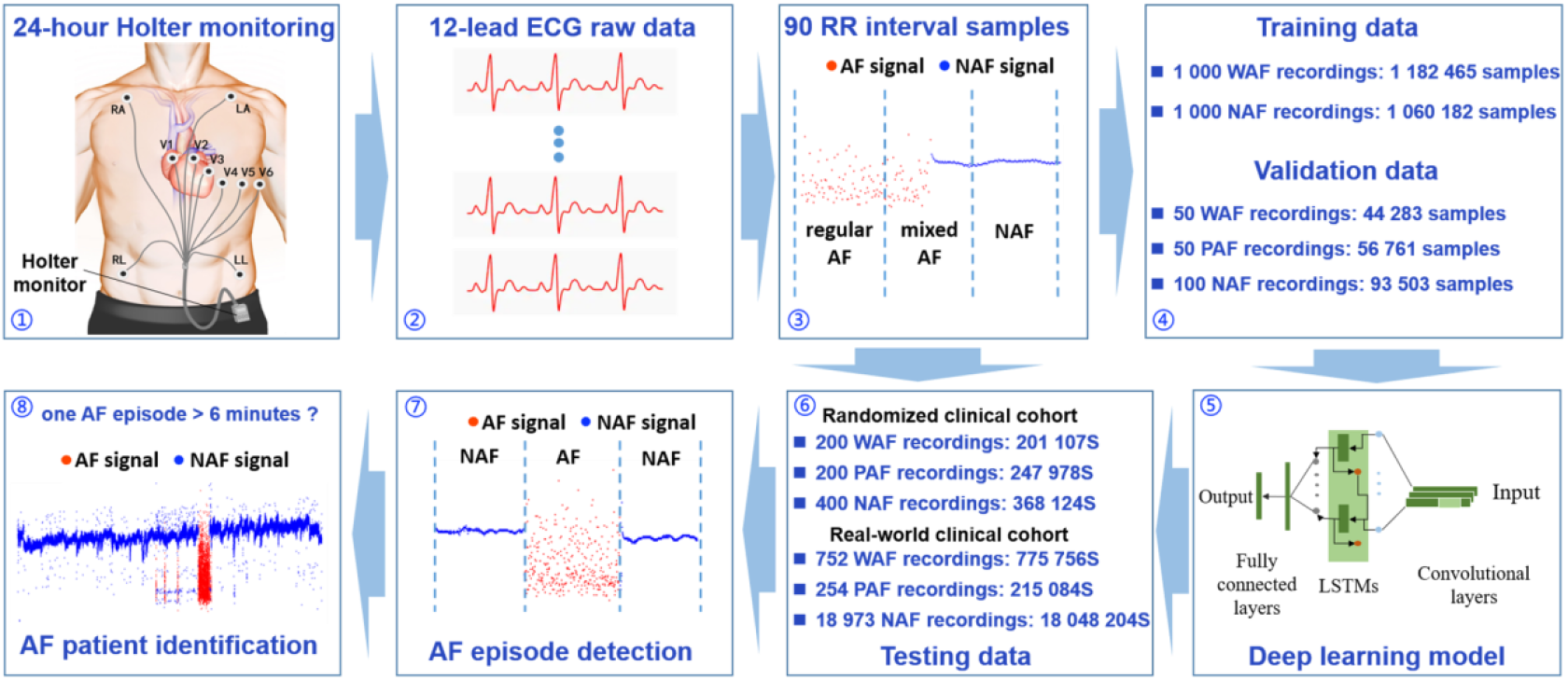
Diagram for automatic atrial fibrillation (AF) episode detection and AF patient identification. The red dots and blue dots represent RR interval of AF signal and non-AF (NAF) signal, respectively. WAF represents whole-course AF. In the composition of testing data, S represents samples. In the AF patient identification, a 24 h recording (RR-interval data) was diagnosed as an AF patient because it contained an AF episode (red dots) longer than 6 min.

### Data sources

Our dataset consisted of 23 408 recordings collected from a total of 23 073 adult patients (age > 18 years) who had a 24 h dynamic 12-lead ECG recording with sampling rate of 512 Hz captured by a Holter machine (DMS Holter Company, Stateline, NV, USA) at three campuses (Main Campus, Optical Valley Campus, and Sino-French New City Campus) of the Cardiac Function Examination Center (Division of Cardiology, Tongji Hospital, Huazhong University of Science and Technology, Wuhan, China). Both the in-hospital and ambulatory patients were pooled together in our dataset. Our dataset included three types of patients: the whole-course AF (WAF), the PAF and the NAF patients. For WAF patient data, whole recording included only AF signals. For NAF patients, the entire recording data had no AF signals but included normal sinus rhythm, sinus arrhythmia, atrial arrhythmia, ventricular arrhythmia, atrioventricular block and so on. The data of PAF patients included both AF episodes and NAF signals. Both WAF and PAF patients were considered to be AF patients. The profile of the dataset is shown in **Figure 2**, our dataset contained the following two cohorts:

**Figure. 2.**
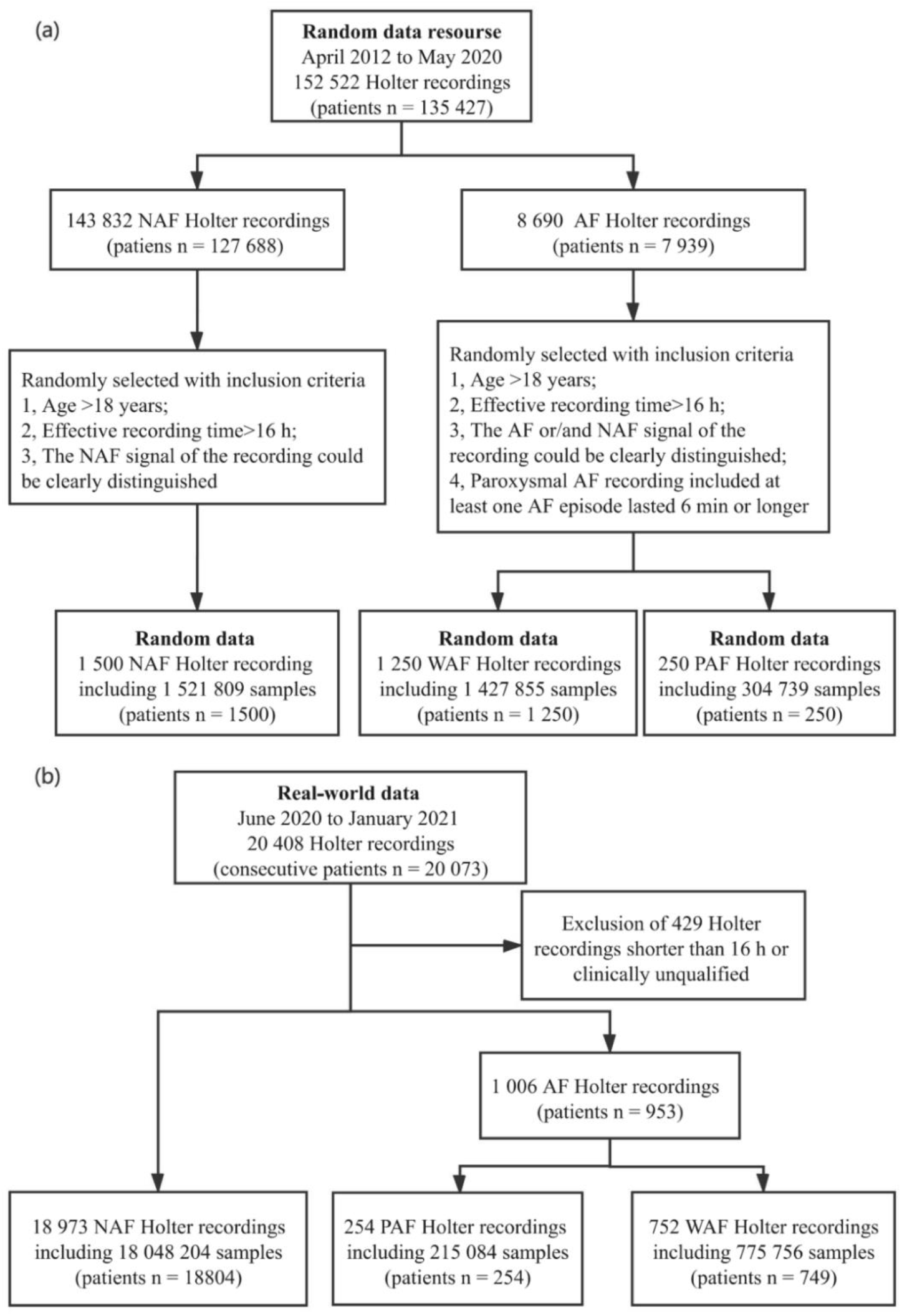
Profile of the datasets. (a) The randomized clinical cohort, and (b) The real-world clinical cohort. PAF = paroxysmal atrial fibrillation (AF) patients; WAF = whole-course AF patients; NAF = non-AF patients.

1. A randomized clinical cohort, including 3 000 recordings from 3 000 adult patients randomly selected from the patient pool enrolled between April 2012 and May 2020. This cohort is employed only for training and preliminarily evaluating the accuracy of the AI model, therefore, strict inclusion and exclusion criteria are used to train a good AI model.
2. A real-world clinical cohort, including 20 408 recordings from all consecutive adult patients (n=20 073) who received a 24 h dynamic 12-lead ECG monitoring in the Cardiac Function Examination Center between June 2020 and January 2021. After the exclusion of 429 recordings shorter than 16 hours or clinically unqualified, 19 979 recordings were used as the test set to assess the real-world clinical value of the AI model. Detailed inclusion/exclusion criteria about the two cohorts are listed in the appendix p 2. The patient characteristics are summarized in Table 1.

**Table 1.** Patient characteristics. Premature atrial contraction (Ventricular premature contraction) >3000 means that there were more than 3000 atrial (ventricular) premature heartbeats during the 24 h Holter monitoring. NAF = non-atrial fibrillation patients; PAF = paroxysmal atrial fibrillation patients; WAF = whole-course atrial fibrillation patients; AVB = atrioventricular block.

|  | **randomized clinical cohort**<br>**(n=3,000)** |  |  | **real-world clinical cohort**<br>**(n=19,979)** |  |  |
| --- | --- | --- | --- | --- | --- | --- |
|  | WAF<br>(n=1,250) | PAF<br>(n=250) | NAF<br>(n=1,500) | WAF<br>(n=752) | PAF<br>(n=254) | NAF<br>(n=18,973) |
| **Age Group** |  |  |  |  |  |  |
| 18 – 25 | 0 | 0 | 52 | 0 | 0 | 383 |
| 26 – 40 | 26 | 3 | 175 | 11 | 7 | 2,235 |
| 41 – 60 | 304 | 74 | 642 | 163 | 61 | 7,596 |
| 61 – 80 | 779 | 152 | 587 | 475 | 158 | 8,102 |
| ≥81 | 141 | 21 | 44 | 103 | 28 | 657 |
| **Sex Group** |  |  |  |  |  |  |
| Male | 777 | 150 | 804 | 471 | 181 | 10,159 |
| Female | 473 | 100 | 696 | 281 | 73 | 8,814 |
| **Pacemaker** |  |  |  |  |  |  |
|  | 1 | 2 | 1 | 28 | 8 | 188 |
| **Premature atrial contraction>3000** |  |  |  |  |  |  |
|  | 0 | 42 | 15 | 0 | 85 | 877 |
| **Ventricular premature contraction>3000** |  |  |  |  |  |  |
|  | 13 | 8 | 18 | 77 | 11 | 961 |
| **First degree AVB** |  |  |  |  |  |  |
|  | 0 | 4 | 43 | 0 | 2 | 577 |
| **Second degree AVB** |  |  |  |  |  |  |
|  | 0 | 1 | 9 | 0 | 0 | 345 |
| **Third degree AVB or complete heart block** |  |  |  |  |  |  |
|  | 3 | 0 | 2 | 4 | 0 | 26 |

The data collection team was responsible for sample collection and anonymization. The algorithm development team received anonymized patient data with new case numbers, age, and gender information for subsequent algorithm development. Written informed consent was not required for this study as the Holter data were collected retrospectively and appropriately anonymized and deidentified according to the Health Insurance Portability and Accountability Act Safe Harbor provision^15^. The study design was evaluated and exempted from full review by the Huazhong University of Science and Technology Institutional Review Board.

### Training, validation, and testing sets

The composition of training, validation and testing data is shown in Figure. 1. The patients in the randomized clinical cohort were randomly divided into training, validation and test sets (10:1:4). Of note, the training data included samples only from WAF patients (positive samples) and NAF patients (negative samples), whereas the validation and testing data also contained samples from PAF patients. In addition to the test set of the randomized clinical cohort, all data in the real-world clinical cohort were also used as the test data to further evaluate the clinical effectiveness of the model. The data in the training set were recorded prior to the data in the real-world clinical cohort; this mimics the real clinical situation in which a model is first trained by existing retrospective data and then tested prospectively on newly acquired data.

### Annotation procedures

All the 24 h ECG data of both cohorts underwent additional annotation procedures. They were initially interpreted by primary cardiologists, then the randomized clinical cohort was further reviewed by three senior board-certified cardiologists, whereas the real-world clinical cohort was reviewed by one senior board-certified cardiologist to ensure the correctness of the base diagnostic labels. The cardiologist committees discussed by consensus the annotated records and provided a reference standard for model evaluation.

Each AF episode included the accurately labeled start time and end time for patients with PAF. The start and end times of each PAF episode were the corresponding time of the first atrial wave with an atrial rate greater than 350 beats/min and the corresponding time of the first P-wave with sinus rhythm after the termination of AF. Specifically, PAF episodes lasting more than 30 s were accurately labeled, and those lasting less than 30 s were labeled as accurately as possible.

Moreover, each interval of premature beat or tachycardia was marked as “A” (atrium event) or “V” (ventricle event) for further labeling six types of arrhythmia with irregular RR intervals: Premature atrial contraction (PAC), Frequent premature atrial contraction (FPAC), Ventricular premature contraction (VPC), Frequent ventricular premature contraction (FVPC), Atrial tachycardia (AT), and Ventricular tachycardia (VT). The long RR interval caused by QRS wave dropping was marked as “B” to further label Second-degree atrioventricular block (AVB2) (appendix p 9). The labels of the data were consistent with the diagnostic results of clinical cardiologists.

### Deep learning model

In this study, we constructed a convolutional, long short-term memory, and fully connected deep neural network (CLDNN) to detect AF episodes automatically. The CLDNN model is composed of three modules, i.e., convolutional neural networks, long short-term memory networks, and deep neural networks (appendix pp 3-4). The 24 h raw ECG data were preprocessed to obtain the 90-RR-interval samples as the input of the model (appendix pp 2-3). For each input of 90-RR-interval sample, the model output a predicted label of AF or NAF. Additional technical details are presented in the appendix pp 3-4.

### Algorithm evaluation

We first detected all AF episodes in 24 h Holter recording data. During the annotation procedure, 90-RR-intervals were labeled as AF or NAF (appendix p 3). Therefore, this detection can be assessed at the level of each 90-RR-interval sample, referred to as “sample-level” results. At this level, the detection result was compared against the reference standard for each 90-RR-interval sample. Based on the “sample-level” results, we further identified whether a patient was an AF patient, referred to as “patient level” results. In “patient level,” a patient was identified as an AF patient if his/her recording included at least one AF episode of more than 6 min.

### Statistical analysis

We used different performance metrics to evaluate the performance of our model, including area under the receiver operating characteristic (ROC) curve (AUC), sensitivity, specificity, and accuracy. Moreover, we used a two-sided 95% confidence interval (CI) to evaluate data variability for each metric^17^. The CI for the AUC was estimated using the Delong method^18^, whereas that for other metrics were obtained using the bootstrap method with 2000 replications. Further, we calculated the ROC curve and the free-response ROC (FROC) curve to further evaluate the detection performance at the “sample-level”. For “patient level” results, the sensitivity and specificity were computed to represent the identification results of different categories of patients.

## Results

In total, 800 recordings (from 800 patients) in the randomized clinical cohort and 19 973 recordings (from 19 739 patients) in the real-world clinical cohort were tested to evaluate the performance of our CLDNN model. The performance was first evaluated at the “sample-level”. In the randomized clinical cohort, 817 209 90-RR-interval samples were tested, and the results are shown in Table 2. Specifically, the model achieved a sensitivity of 0.982 and specificity of 0.993 for testing samples in PAF patients, and correctly detected 99.7% of the AF samples in WAF patients and 99.9% of the NAF samples in NAF patients.

**Table 2.**
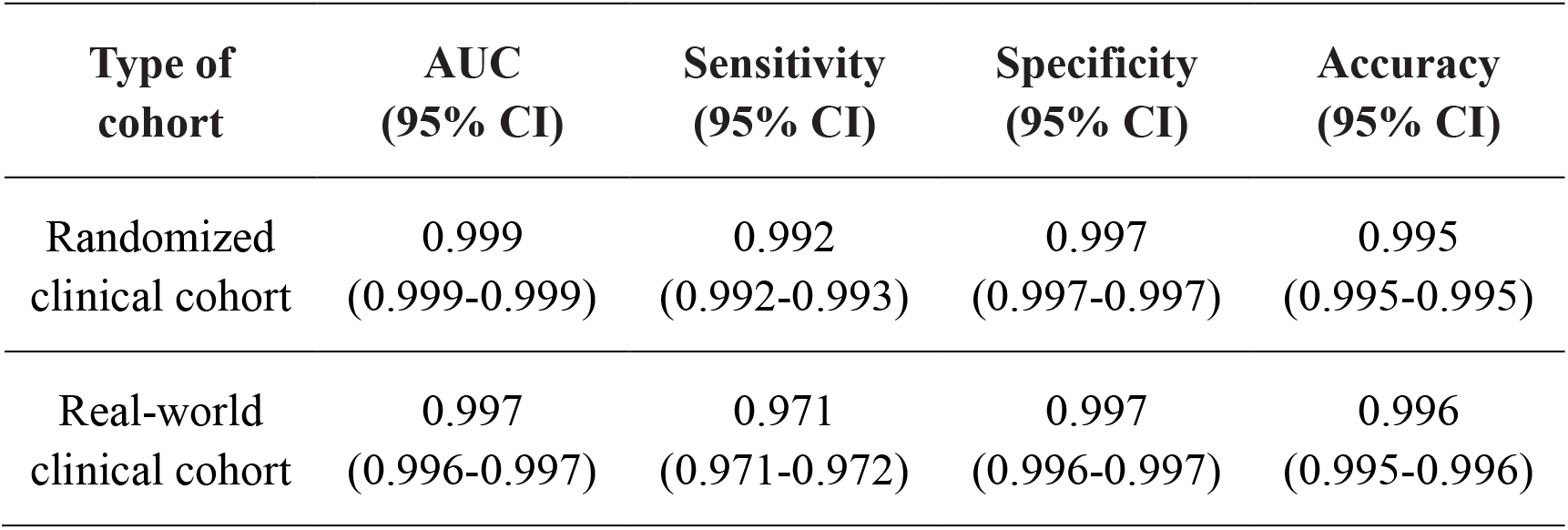
AF detection performance at the “sample-level” for all patients. The AF episode detection performance was evaluated for both cohorts. AF = atrial fibrillation; AUC = area under the curve; CI = confidence interval.

We further analyzed the AF detection performance in PAF patients with different proportions of AF episodes (appendix p 17), the results are summarized in appendix p 11. In general, the model achieved good performance for all groups with an AUC greater than 0.98. However, when the proportion of AF episodes were lower than 10%, the sensitivity was decreased to 0.872; the performance was improved as the proportion of AF episodes increased.

In the real-world clinical cohort, 19 039 044 samples were tested to further evaluate the clinical effectiveness of the AI model, and the results are shown in Table 2. Specifically, the model achieved a sensitivity of 0.964 and specificity of 0.987 for testing samples in PAF patients, and correctly detected 97.3% of the AF samples in WAF patients and 99.7% of the NAF samples in NAF patients.

Moreover, Figure. 3 shows the ROC curve and FROC curve for the “sample-level” analyses of all the testing recordings in both cohorts. For the ROC curves, our model achieved an AUC of 0.997 (95% CI 0.996–0.997) and 0.999 (95% CI 0.999–0.999) for the real-world clinical cohort and randomized clinical cohort, respectively. For the FROC curves, at a rate of one false positive sample per recording, our model achieved a sensitivity of 0.950 and 0.980 in the real-world clinical cohort and randomized clinical cohort, respectively. In general, our model achieved very good performance for both cohorts, and the performance of the model decreased slightly in the real-world clinical cohort. In addition, our method obtained superior and consistent performance in the external public database (appendix p 6).

**Figure. 3.**
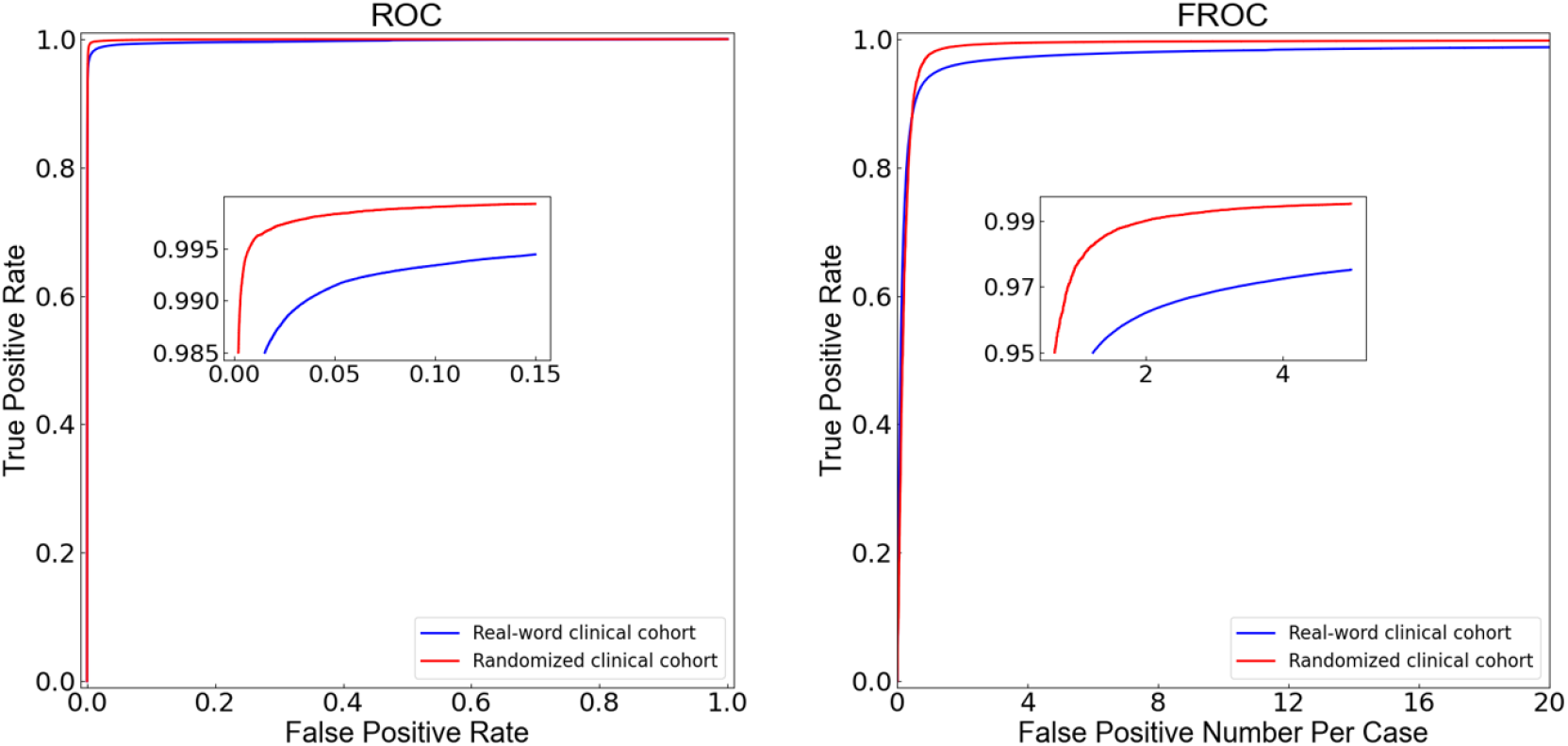
The ROC curve and FROC curve for the “sample-level” analyses in the two cohorts. ROC = receiver operating characteristic. FROC = free-response receiver operating characteristic.

Based on the above results of “sample-level”, the performance of “patient level” was further evaluated for both cohorts, and the results are shown in Table 3. For the randomized clinical cohort, all the WAF and 98.5% of the PAF patients were correctly identified as AF patients, and 99.0% of the NAF patients were correctly identified as NAF patients. For the real-world clinical cohort, 99.3% of WAF and 100.0% of PAF patients were correctly identified as AF patients, and 97.4% of the NAF patients were correctly identified as NAF patients. The ROC curves for the “patient level” analyses in the two cohorts are shown in Figure. 4.

**Table 3.** AF patient identification performance at the “patient level”. The performance of AF patient identification was evaluated for both cohorts. AUC = area under the curve; CI = confidence interval.

| Type of cohort | AUC (95% CI) | Sensitivity (95% CI) | Specificity (95% CI) | Accuracy (95% CI) |
| --- | --- | --- | --- | --- |
| Randomized clinical cohort | 0.998 (0.998-0.998) | 0.993 (0.987-1.000) | 0.990 (0.980-0.997) | 0.991 (0.986-0.997) |
| Real-world clinical cohort | 0.997 (0.997-0.997) | 0.995 (0.989-0.999) | 0.974 (0.970-0.975) | 0.975 (0.972-0.976) |

**Figure. 4.**
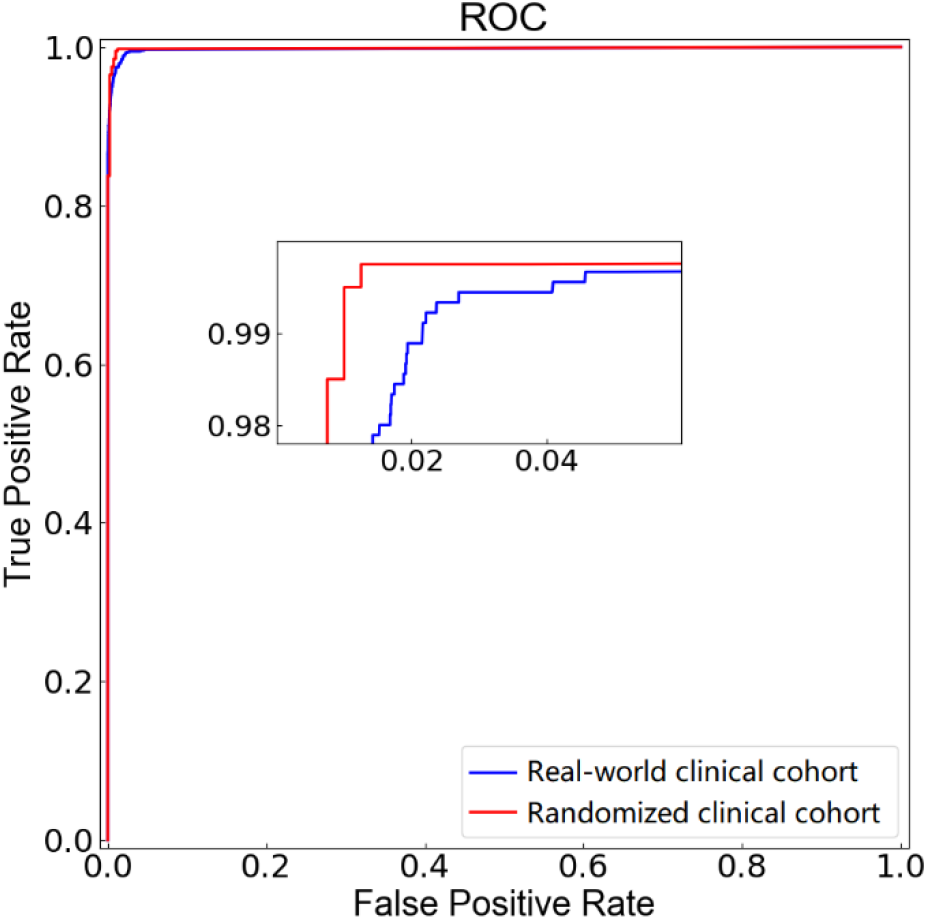
The ROC curve for the “patient level” analyses in the two cohorts. ROC = receiver operating characteristic.

RR-interval-based approach faces difficulty in distinction between AF and some other arrhythmia with irregular RR intervals. This difficulty was concerned in many studies; however, it has not been systematically and quantitatively studied^19,20^. Based on the above results of “sample-level” analyses, we further quantitatively evaluated the performance of our model in distinguishing AF from other arrhythmias with irregular RR intervals in the NAF recordings of both cohorts. Seven types of arrhythmia with irregular RR intervals were included and the detection results at the “sample-level” are shown in Table 4. In general, our model achieved comparable performance for both cohorts. Our model achieved an average accuracy of 0.995 in specifically detecting the NAF sample in all the seven types of arrhythmia for the randomized clinical cohort. For the real-world clinical cohort, the average accuracy was 0.985, with a relatively lower accuracy of 0.854 for AT.

**Table 4.** The results of distinguishing AF from other arrhythmias with irregular RR interval. Seven types of arrhythmia with irregular RR interval were included: Premature atrial contraction (PAC), Frequent premature atrial contraction (FPAC), Ventricular premature contraction (VPC), Frequent ventricular premature contraction (FVPC), Atrial tachycardia (AT), Ventricular tachycardia (VT), and Second-degree atrioventricular block (AVB2).

|  | **Randomized clinical cohort**<br>**(Sensitivity:0.992)** |  | **Real-world clinical cohort**<br>**(Sensitivity:0.971)** |  |
| --- | --- | --- | --- | --- |
| **Type** | **Number of**<br>**samples** | **Specificity**<br>**(95% CI)** | **Number of**<br>**samples** | **Specificity**<br>**(95% CI)** |
| FPAC | 5,668 | 0.989<br>(0.987-0.992) | 392,875 | 0.966<br>(0.965-0.966) |
| PAC | 14,950 | 1.000<br>(0.999-1.000) | 1,099,795 | 0.995<br>(0.995-0.995) |
| FVPC | 9,777 | 0.999<br>(0.998-0.999) | 502,957 | 0.992<br>(0.992-0.992) |
| VPC | 17,348 | 0.999<br>(0.999-1.000) | 972,030 | 0.995<br>(0.995-0.995) |
| AT | 1,909 | 0.913<br>(0.900-0.926) | 109,723 | 0.854<br>(0.852-0.826) |
| VT | 103 | 1.000<br>(1.000-1.000) | 9,855 | 0.972<br>(0.969-0.975) |
| AVB2 | 2 | 1.000<br>(1.000-1.000) | 28,598 | 0.902<br>(0.899-0.906) |
| Other NAF | 321,038 | 0.997<br>(0.996-0.997) | 15,156,192 | 0.999<br>(0.999-0.999) |
| Average of<br>Seven types | 49,757 | 0.995<br>(0.994-0.995) | 3,115,833 | 0.985<br>(0.985-0.985) |
| All | 368,124 | 0.997<br>(0.997-0.997) | 18,048,204 | 0.997<br>(0.996-0.997) |

Finally, we analyzed the misclassified cases in “patient-level” for real-world clinical cohort, the results are shown in Table 5. A total of five cases of WAF were misclassified as NAF patients, including one case of complete heart block with AF, four cases of complete ventricular pacing with AF. A total of 501 NAF patients were misclassified as AF, including sick sinus syndrome (SSS), atrial high-rate episode (AHREs), PAC, VPC, AVB and other cases.

**Table 5:** The analysis of misclassified cases in “patient level” for real-world clinical cohort.

| **Types of arrhythmia** | **False negative patients**<br>**(n=5)** |
| --- | --- |
| Complete heart block with AF | 1 |
| AF with complete ventricular pacing | 4 |
|  | **False positive patients**<br>**(n=501)** |
| Sick sinus syndrome | 75 |
| Atrial high-rate episode | 68 |
| Premature atrial contraction | 112 |
| Ventricular premature contraction | 25 |
| Atrioventricular block | 11 |
| Others | 210 |

## Discussion

In this study, we developed an AI pipeline for automatic AF episode detection and AF patient identification in 24 h Holter recording. To the best of our knowledge, this is the first study that automatically identifies both AF episodes and AF patients in 24 h Holter recordings, and achieved high accuracy in testing with the large real-world clinical dataset.

This study used the RR intervals as the input to the deep learning model for two reasons: 1) The irregularity of RR intervals is a main feature of the AF^21^;2) Another feature of the AF is the appearance of fine or coarse fibrillatory waves and the absence of P-wave^22^, however, this feature can be easily confused by common perturbations in Holter recording^23^. Our experiments showed that our RR-interval-based method and the raw-ECG-based method achieved similar performance (appendix p 4-6), which indicates that the RR-interval-based features may be the major ones for detecting AF in the deep learning model. Our experiments further showed that the RR-interval-based method can distinguish (with an average accuracy of 0.985) AF from seven types of other arrhythmia with irregular RR intervals, which are commonly considered to be difficult to recognize by use of RR intervals^19,20^. Finally, in comparison with the state-of-the-art methods using a public dataset (appendix p 6), our method obtained consistent and superior performance, which further proved the effectiveness of our RR-interval-based method in AF detection.

Compared with the raw-ECG-based method (appendix pp 4-6), the computational cost of RR-interval-based method was significantly lower. Our RR-interval-based method could process a 24 h Holter recording within 5 s (appendix pp 6-7), while the raw-ECG-based method used approximately 2 min. In terms of storage space, the size of each raw ECG recording (>200 MB) is approximately 50 times larger than that of RR-interval data of each recording (<4 MB). In addition, a recent study published in *Nature Medicine* showed that the common perturbations (adversarial attack) to single-lead ECG could lead to a 74% misdiagnosis rate for raw-ECG-based deep learning model. In contrast, RR-interval-based method seemed more robust to such adversarial attacks^24^. Moreover, another advantage of using RR interval (heartbeat) rather than raw ECG data is its independence to the ECG waveform^21^. Our method can be widely applied to medical devices that can collect heartbeat information, including not only the multi-lead and single-lead Holter devices, but also other wearable devices that can reliably measure the heartbeat signals.

Few studies have used long-term Holter recording data for automatic AF detection^25^; it is a remarkable challenge to accurately label long-term Holter monitoring ECG data of many PAF patients. In this study, we used AF samples from WAF patients, instead of PAF patients, to train the AI-model, thus providing a strategy to circumvent the challenge. The 24 h Holter ECG data from WAF patients are significantly easier to label than the data from PAF patients. High detection performance in this study indicated that it was feasible to accurately detect the PAF samples using a model trained with WAF samples. This is the first study that trained the AI-model with WAF samples, and tested with the PAF samples. Moreover, we used data of 2 000 recordings to train our AI model and we also tested the effect of training dataset size on the detection performance (appendix p 6). It appeared that the training dataset of 2,000 recordings was considered sufficient to achieve robust performance in automatic AF episode detection and AF patient identification.

At present, there is no well-recognized criterion for automatically identifying AF patient in long-term Holter monitoring. Even if the automatic detection of AF episodes can achieve good performance in applying to AF screening, clinician intervention will still be needed to identify patients with AF, because correctly identifying all segments of the 24 h Holter recording is an unattainable goal. In the absence of reliable criterion to determine the diagnostic ability at the “patient level”, further detection and over-treatment may cause injury while clinical time and medical resources may be wasted. In this study, we identified AF patients by using a criterion of at least one AF episode of 6 min or longer, as this condition significantly increased the risk of thromboembolism (stroke, transient ischemic attack, or systemic embolism)^26,27^. We also tested other thresholds of AF episode in AF patient identification and found that the threshold of 6 min achieved the best performance (appendix p 12). Our criterion not only achieved good performance but also had clinical interpretation, therefore would be a good criterion for automatic identification of AF patients in clinical application.

The data used in most AI-related clinical studies are selected with strict inclusion criteria, and the marginal or uncertain data are excluded, which leads to the decline of model performance in real clinical application^28^. To minimize selection bias, our model has been further verified by the real-world data, and achieved consistent good performance while exposes shadow spots for some special data. The expert committee re-analyzed these data at the “sample-level” and “patient level”. At the “sample-level”, the samples with AHREs and AVB2 complicated with sleep apnea syndrome were very similar with samples of AF, which were easily misclassified as AF. However both of them have a high risk of AF^27^, and such “misclassifications” may have positive meaning for prevention of AF. At the “patient level”, AF with AVB3 and slow ventricular rate AF with ventricular pacing were easily judged as NAF, because they had regular rhythms. More analysis of the misclassifications is presented in appendix p 8. In general, the analysis of misclassifications in real-world clinical data help us find shadow spots of our model in the detection of some rare and special ECG signals. According to the shadow spots, the application range can be set in advance, and the model can be further improved in the later stage. Therefore, real-world data testing is an important step in the clinical application of deep learning models.

Our study has a few limitations. First, the mixed AF samples (including both AF and NAF RR-intervals) could not be detected as accurately as regular AF samples (including only AF RR-intervals), which led to relatively lower AF detection performance in PAF patients than in WAF or NAF patients. We believe that this limitation had limited effect on the overall performance of AF detection because the mixed AF samples only accounted for a very small part of a 24 h Holter recording. The second limitation is that our method used a criterion of at least one AF episode of 6 min or longer to identify AF patients, therefore, it was not able to identify the AF patient with episode of less than 6 min. Finally, our CLDNN model achieved relatively lower accuracy in distinguishing AF from the 7 types of arrhythmia with irregular RR intervals. A more effective AI-model is needed to improve the detection accuracy for these 7 types of arrhythmias.

In conclusion, we developed a deep learning model to automatically detect AF episode and diagnose AF patients in long-term Holter recordings. Our method executed very fast (in a few seconds) and achieved high specificity and sensitivity in both AF episode detection and AF patient identification for a large real-world clinical dataset. Therefore, it has great potential to help clinicians improve AF diagnostic accuracy and reduce workload, which will promote AF screening with lower cost to effectively reduce the risk of stroke caused by AF.

## Data Availability

Python scripts for data preprocessing and modelling can be requested by contacting corresponding author QL. The test dataset of the randomized clinical cohort in this study is publicly available at Mendeley Data, V1, DOI:10.17632/6jd4rn2z9x.1. The MIT-BIH atrial fibrillation database is a public dataset available at: https://archive.physionet.org/physiobank/database/afdb/. The training and validation sets in the randomized clinical cohort and all the data in the real-world clinical cohort will be available after approval by the corresponding author XY. Any data and code use will be restricted to non-commercial research purposes.

https://data.mendeley.com/datasets/44htzjcgsz/1

## Contributors

PZ, FL, XY and QL were responsible for the study design. DW, XY and QL conceived the project. PZ, YC and QL chose the architecture, implemented and tuned the deep neural network. PZ and YC did the statistical analysis of the test data and generated the figures and tables. FL, FM, XY and DW were responsible for collecting, preprocessing and annotating the datasets. PZ, FL, FM and QL contributed to the writing and all authors revised it critically for important intellectual content. PZ, FL, XY and QL have directly accessed and verified the underlying data reported in the manuscript. All authors read and approved the submitted manuscript.

## Data sharing

Python scripts for data preprocessing and modelling can be requested by contacting corresponding author QL. The test dataset of the randomized clinical cohort in this study is publicly available at Mendeley Data, V1, doi: 10.17632/44htzjcgsz.1. The MIT-BIH atrial fibrillation database is a public dataset available at: https://archive.physionet.org/physiobank/database/afdb/. The training and validation sets in the randomized clinical cohort and all the data in the real-world clinical cohort will be available after approval by the corresponding author XY. Any data and code use will be restricted to non-commercial research purposes.

## Acknowledgments

The authors sincerely thank Mr. Fang Yu of DMS Holter Company for his technical support and Mrs. Zhiying Zhang for her support in manuscript editing. This research was funded by the National Natural Science Foundation of China (Grant No. 62006087, 81500328), Science Fund for Creative Research Group of China (Grant No. 61721092), Director Fund of Wuhan National Laboratory for Optoelectronics (WNLO).

## SUPPLEMENTARY APPENDIX

### Supplementary Methods

#### 1. Detailed inclusion/exclusion criteria of the two cohorts

For the randomized clinical cohort used for training and preliminary testing of the artificial intelligence (AI) model, the inclusion criteria were: 1) the patients must be 18 years or older; 2) the atrial fibrillation (AF) or/and non-AF (NAF) signal of the patients could be clearly distinguished; 3) the effective recording time was longer than 16 h; and 4) the data of paroxysmal AF (PAF) patients included the sinus rhythm and at least one AF episode lasted 6 min or longer, because the AF patients were identified based on a 6 min duration of AF episode in this study. Moreover, this cohort contained the same number of the AF and NAF patients both in the training and testing sets, and the AF patients included the same number of the whole-course AF (WAF, n=200) and PAF (n=200) patients in the testing set. Therefore, this cohort can be used to well evaluate the detection performance of the AI model for various types of patients.

Different from the randomized clinical cohort, the real-world clinical cohort included all consecutive adult patients, except those with shorter than 16 hours of recording time or those with clinically unqualified recording (n=429). There were a total of 254 PAF, 752 WAF and 18 973 NAF qualified recordings. This real-world clinical cohort can be used to well evaluate the clinical practicability of the AI model.

It is worthwhile to mention two special groups in the 254 PAF recordings. The first group included PAF recordings (n=54) in which AF episodes were less than 6 min. These recordings could still be tested at the “sample-level”, but not at the “patient level”, because the AF patients were identified based on a 6 min duration of AF episode. The second group included PAF patients (n=53) whose recordings contained AF and NAF signals that were too ambiguous to clearly distinguished by the clinicians. These recordings could still be tested at the “patient level”, but not at the “sample-level”.

#### 2. Data preprocessing

Raw ECG signals were preprocessed using manufacturer-specific commercial software for the DMS CardioSca12 Satellite System (DMS Holter Company, Stateline, NV, USA) to obtain the RR interval data. Noise was automatically detected and marked by this commercial software. For the preprocessed RR interval data, we used a nonoverlap sliding window with a length of 90-RR-interval to extract the RR interval samples; samples with detected noise were removed. Significantly, there were still a lot of noise that could not be detected by the commercial software in the samples. The AI model in this study was trained and evaluated based on these RR interval samples. The training data included samples only from WAF patients (positive samples) and NAF patients (negative samples), whereas the validation and testing data also contained samples from PAF patients.

Each 90-RR-interval sample was labeled as either an AF (positive) or a NAF (negative) sample according to the labeled start and end times of the AF episodes. Samples extracted from WAF patients were all AF, and those from NAF patients were all NAF. Some testing samples extracted from the PAF patients contained both AF and NAF RR intervals; these samples were also labeled as AF. For clarity, however, they were referred to as mixed AF samples for distinction from regular AF samples in the following analysis. To evaluate the performance of our model in distinguishing AF from other arrhythmias that also have the irregular RR intervals, each 90-RR-interval sample of the NAF patients was further labeled as: FPAC, PAC, FVPC, VPC, AT, VT, AVB2 or other NAF sample using the standard described in Table S1 based on the annotation of “A”, “B” or “V”. Because a 90-RR-interval sample might contain “A”, “B”, “V” at the same time, the sample could have multiple labels and was evaluated independently for each label in testing.

#### 3. Additional technical details of the deep learning model

##### 3.1 The architecture of the deep learning model

The architecture of the model is illustrated in Figure. S1 and the model is composed of three modules, i.e., convolutional neural networks (CNNs), long short-term memory networks (LSTMs), and deep neural networks (DNNs). Specifically, the CNNs were composed of two convolutional layers with 64 kernels of size 5 and 32 kernels of size 3, respectively. We tested the implementation of batch normalization and max pooling in the CNN module, but found no improvement. After the convolutional layers, we used bidirectional LSTM to utilize the forward and backward information of the input. We took the output of all time steps as the output of bidirectional LSTM and used them as the input of the global max pooling layer. The DNN module consisted of two fully connected layers. Before each fully connected layer, we applied dropout with a probability of 0.2 to prevent over-fitting and improve the generalization ability. The final fully connected softmax layer, as the output of our model, produced a distribution over AF and NAF. The ReLU activation function was applied after each layer, except the output layer.

##### 3.2 Model training and hyperparameter tuning

The model weights were randomly initialized with a Glorot uniform, while the bias was initialized with zeros. We trained the model using the categorical cross-entropy loss function with the Adam optimizer, taking the default parameters β_1_=0.9 and β_2_=0.999, and initialized the learning rate to 1e-3. The model was trained with a batch size of 128 at each epoch, and the learning rate was reduced by a factor of 0.1 when the training loss stopped improving for three consecutive epochs.

In general, the hyperparameters of our model were determined via grid search and manual tuning. We obtained the final hyperparameters with the following steps: (i) selected the model parameters in the training stage and (ii) checked the performance in the validation stage. Specifically, the hyperparameters were selected from the following options: convolutional layer filters [16, 32, 64, 96], kernel size [3, 5, 7], batch size [16, 32, 64, 128], and dropout rate [0.1, 0.2, 0.3]. The actual performance of the model was important to guide us in selecting the hyperparameters. During the manual tuning procedure, we determined the next parameter selection based on the performance of previous iterations. In addition, we modified the architecture of the original CLDNN to fit our task. We also attempted to remove some unnecessary structures and connections, but increased other modules with performance improvement.

For the trained model, when a new 90-RR-interval sample was input, the output would be a predicted label to indicate if the input sample was AF or not. After all samples from a 24 h Holter recording were tested, we further employed a “correction” strategy to improve the detection accuracy; if an isolated sample was predicted as AF (NAF) among four or more consecutive NAF (AF) samples, it was corrected as NAF (AF).

#### 4. Raw ECG-based method for AF detection

For the raw ECG-based method, we used the same training, validation and testing data of the randomized cohort as our RR interval-based method did. The raw ECG data were exported with sampling rate of 128 Hz, and were filtered using an elliptical band-pass filter with the cutoff frequency of 0.5 Hz and 35 Hz to remove baseline wander and high frequency components. Each raw recording with 12 leads was then divided into segments of 8 seconds and each segment was the dimension of 1 024 * 12; approximately 10 000 samples were extracted from each patient. The sample size of the raw data is too large to be processed by our equipment, therefore, we randomly selected 100 samples from each patient of the training and validation sets, and obtained a total of 200 000 training samples and 20 000 validation samples. All 7 701 706 testing samples from the testing set were used for evaluation.

We used a convolutional DNN as proposed in the previous study^1^ as the model of raw-ECG-based method, and the source convolutional DNN was modified to fit our data. This model had 35 layers which were composed of a convolutional layer, 16 residual blocks (each block included two convolutional layers), and two fully connected layers. The convolutional layer had a filter width of 32 at start and doubled every four residual blocks. The length of the input sample was reduced by half using max pooling after every two residual blocks. The batch normalization and ReLU activation were applied before each convolutional layer, and the dropout layer with a probability of 0.2 was also applied after nonlinearity. The final fully connected softmax layer produced the distribution of AF and NAF.

The model weights were randomly initialized with he normal, and the bias was initialized with zeros. The Adam optimizer with the default parameters β_1_ = 0.9 and β_2_ = 0.999 was used to update the model with the categorical cross entropy loss function, and the learning rate was initialized to 1e-3. We trained the model with a batch size of 128 and 30 epochs, and the learning rate was reduced by a factor of 0.1 when the validation loss stopped improving for five consecutive epochs. Moreover, we chose the hyper-parameters of the model and optimization algorithm via a combination of grid search and manual tuning. Specifically, we primarily optimized the size of the convolutional filters, the number of the final fully connected layers, as well as the number of residual blocks.

### Supplementary Results

#### 1. The AI model performance in public database

We compared the performance of our method with the state-of-the-art methods using the MIT-BIH AF database that is the most commonly used long-term ECG public dataset. This dataset mainly includes 23 records from 23 different patients and each record is approximately 10 h. For fairness concerning, all selected methods used subject-wise independent testing with individual results and cross validation to obtain the sensitivity, specificity and accuracy, the results are shown in Table S2.

#### 2. The performance of AF detection using raw ECG data

The raw ECG-based method employed the same training, validation and test sets of the randomized clinical cohort, as our RR interval-based method did. Figure. S2 shows the AF detection performance for all patients obtained with the raw ECG- and RR-interval-based methods at both the “sample-level” and “patient level”. At the “sample-level”, the raw-ECG-based method achieved a sensitivity of 0.987 and specificity of 0.988 for testing samples from PAF patients, correctly detected 98.6% of the AF samples in WAF patients, and 99.5% of the NAF samples in NAF patients. In “patient level”, 100% WAF patients, 98% PAF patients and 99% NAF patients were correctly identified. In general, our RR-interval-based method achieved slightly higher performance than the raw-ECG-based method at both the “sample-level” and “patient level”. The receiver operating characteristic (ROC) curves for the two methods at the “sample-level” and “patient level” are shown in Figure. S3.

#### 3. Effect of training dataset size on our AI model

In this study, we trained the AI model with 24 h Holter recording data from 1 000 WAF patients and 1 000 NAF patients. To examine the effect of the training dataset size on the detection performance, training datasets with different sizes were used to train the model, whereas the testing dataset remained unchanged. This effect was examined with the randomized clinical cohort. We calculated the ROC curves in Fig. S4 for both the “sample-level” and “patient level” analyses with different training dataset sizes. Starting with a smaller training dataset, an increase in the training dataset improved the AF detection performance in both the “sample-level” and “patient level” analyses. However, when the training dataset size reached 1 500 patients, the AF detection performance no longer improved. Therefore, the training dataset of 2 000 patients was considered sufficient in this study to achieve robust performance in automatic AF episode detection and AF patient identification.

#### 4. Execution time of our method

The time cost is another important aspect in evaluating clinical practicality of a method. The execution time of our method included the time to extract 90-RR-interval samples, to detect the AF episodes, and to identify AF patient given a 24 h Holter recording. We calculated the execution time for cases in both cohorts, the execution time was 4.28±1.92 s for data of WAF patients, 4.47±1.59 s for data of PAF patients, and 3.26±1.02 s for data of NAF patients. On average, our method can process a 24 h Holter recording within 5 s and thus is practical in clinical application.

### Supplementary Discussion

#### 1. Analysis of the misclassifications

In distinguishing AF from 7 types of arrhythmia with irregular RR intervals in the real-world clinical cohort, our results showed that the accuracies were relatively lower for the AT (accuracy of 0.854) and AVB2 (accuracy of 0.902). This may be because the change of RR interval in some samples of the AT or AVB2 were very similar with that in AF, such as the samples with atrial high-rate episodes (AHREs) or with AVB2 complicated with sleep apnea syndrome. AHREs (including atrial flutter) are classified as subclinical AF, and sleep apnea syndrome is a high risk of AF. Therefore, these samples were misclassified as AF because they did have a high risk of AF, and such “misclassifications” may have positive meaning for prevention of AF, instead of simply classifying them as NAF.

In the misclassified cases in Table 5, five cases (one complete heart block with AF, four complete ventricular pacing cases with AF) were misclassified as NAF patients. Unlike other cases of AF, the RR interval was nearly constant throughout the whole recordings in these five cases, which led to these false negatives. The false positive cases mainly included sick sinus syndrome (SSS), AHRE, premature atrial contraction (PAC) and ventricular premature contraction (VPC, the PAC and VPC are mainly of parasystolic rhythm types) whose rhythms had similar RR intervals to the AF and thus can easily lead to these false positives. Moreover, both AHRE and SSS are considered to be causes of stroke, and have a higher risk of AF.

## Supplementary Tables

**Table S1.** The definition of the seven types of arrhythmia with irregular RR interval. The seven types of arrhythmia with irregular RR interval were Premature atrial contraction (PAC), Frequent premature atrial contraction (FPAC), Ventricular premature contraction (VPC), Frequent ventricular premature contraction (FVPC), Atrial tachycardia (AT), Ventricular tachycardia (VT), and Second-degree atrioventricular block (AVB2).

| Type of arrhythmia | Definition |
| --- | --- |
| FPAC | The number of 'A' $\geq 6$ in the 90-RR-interval |
| PAC | The number of 'A' $\geq 1$ and $< 6$ in the 90-RR-interval |
| FVPC | The number of 'V' $\geq 6$ in the 90-RR-interval |
| VPC | The number of 'V' $\geq 1$ and $< 6$ in the 90-RR-interval |
| AT | The number of continue 'A' $\geq 3$ in the 90-RR-interval |
| VT | The number of continue 'V' $\geq 3$ in the 90-RR-interval |
| AVB2 | The number of 'B' $\geq 1$ in the 90-RR-interval |

**Table S2.**
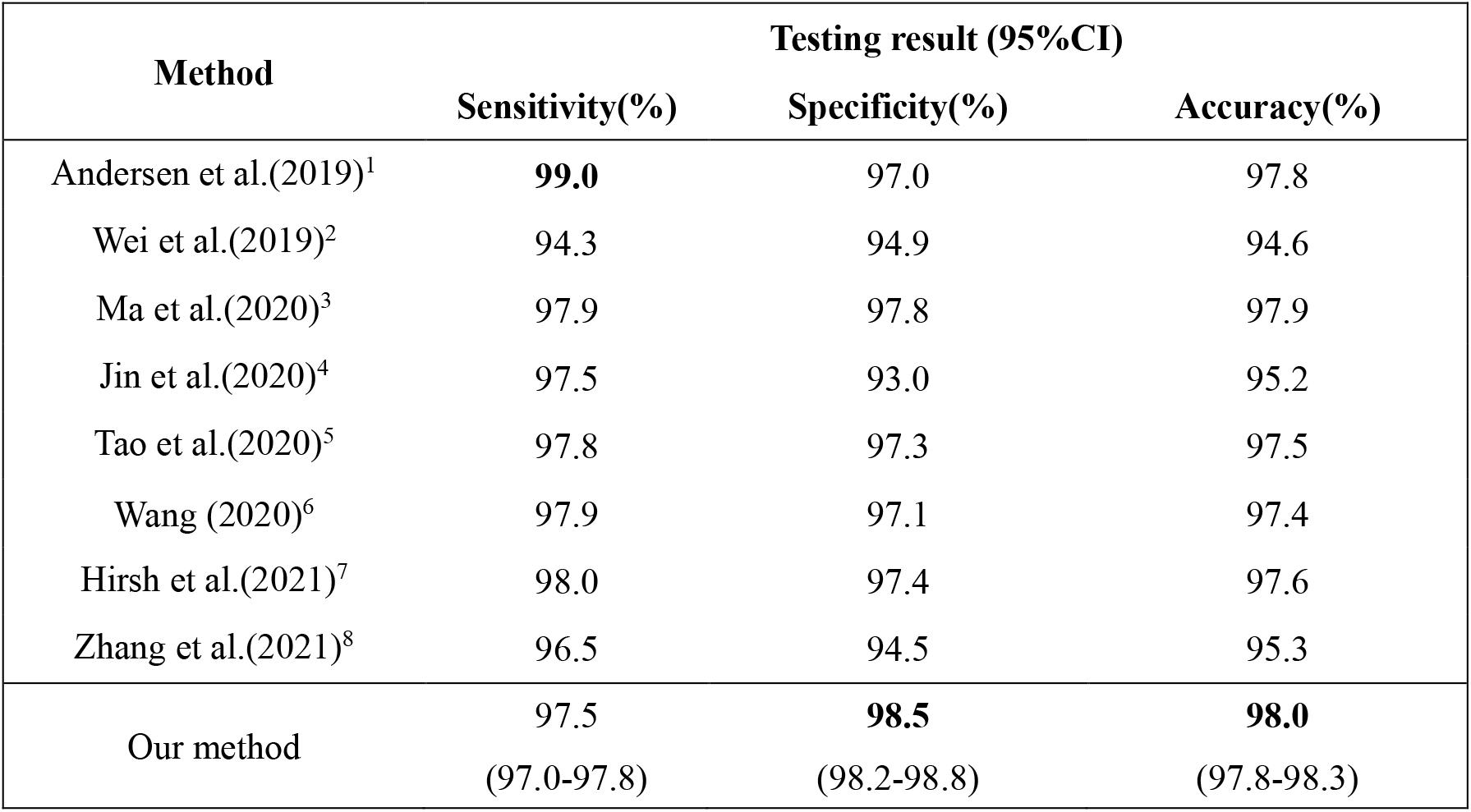
Comparison of atrial fibrillation detection performance of our method and the state-of-the-art methods. CI = confidence interval.

---

**Table S3.**
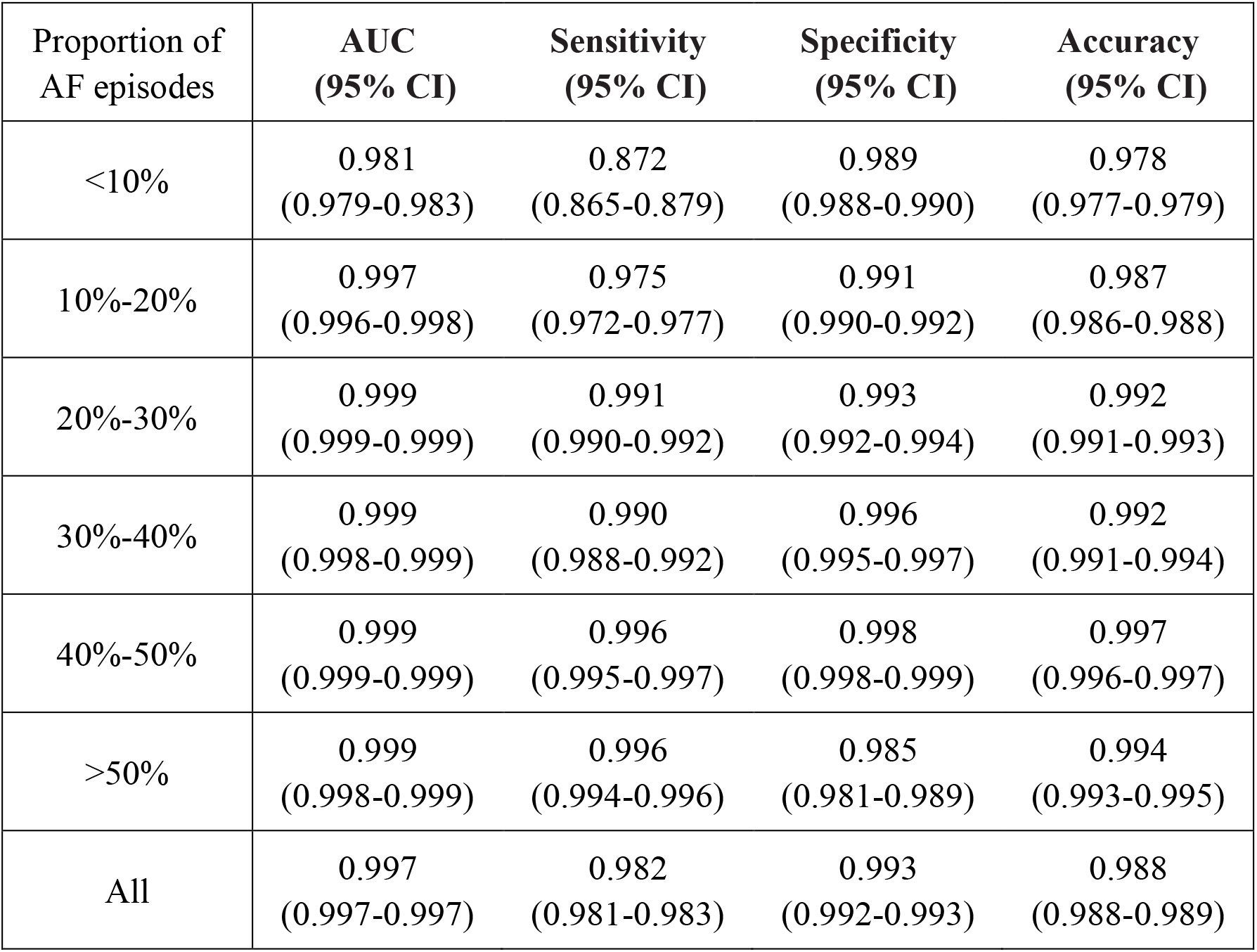
AF detection performance at the “sample-level” for paroxysmal AF patients with different proportion of AF episode. AF = atrial fibrillation; AUC = area under the receiver operating characteristic curve. CI = confidence interval.

**Table S4.** AF detection performance at the “patient level” using different thresholds of AF episode in the randomized clinical cohort. Four thresholds of AF episode were tested in AF patient identification, the threshold of 6 min and 8 min achieved the same performance. With the same accuracy, the shorter threshold may have advantages, therefore, the threshold of 6 min was selected. CI = confidence interval.

| **Threshold**<br>**(min)** | **Sensitivity**<br>**(95% CI)** | **Specificity**<br>**(95% CI)** | **Accuracy**<br>**(95% CI)** |
| --- | --- | --- | --- |
| 3 | 0.998 | 0.973 | 0.985 |
|  | 0.992-1.000 | 0.956-0.988 | 0.976-0.993 |
| 6 | 0.993 | 0.990 | 0.991 |
|  | 0.987-1.000 | 0.980-0.997 | 0.986-0.997 |
| 8 | 0.993 | 0.990 | 0.991 |
|  | 0.983-1.000 | 0.980-0.997 | 0.985-0.997 |
| 10 | 0.988 | 0.990 | 0.989 |
|  | 0.976-0.998 | 0.980-0.997 | 0.981-0.995 |

## Supplementary Figures

**Figure S1.**
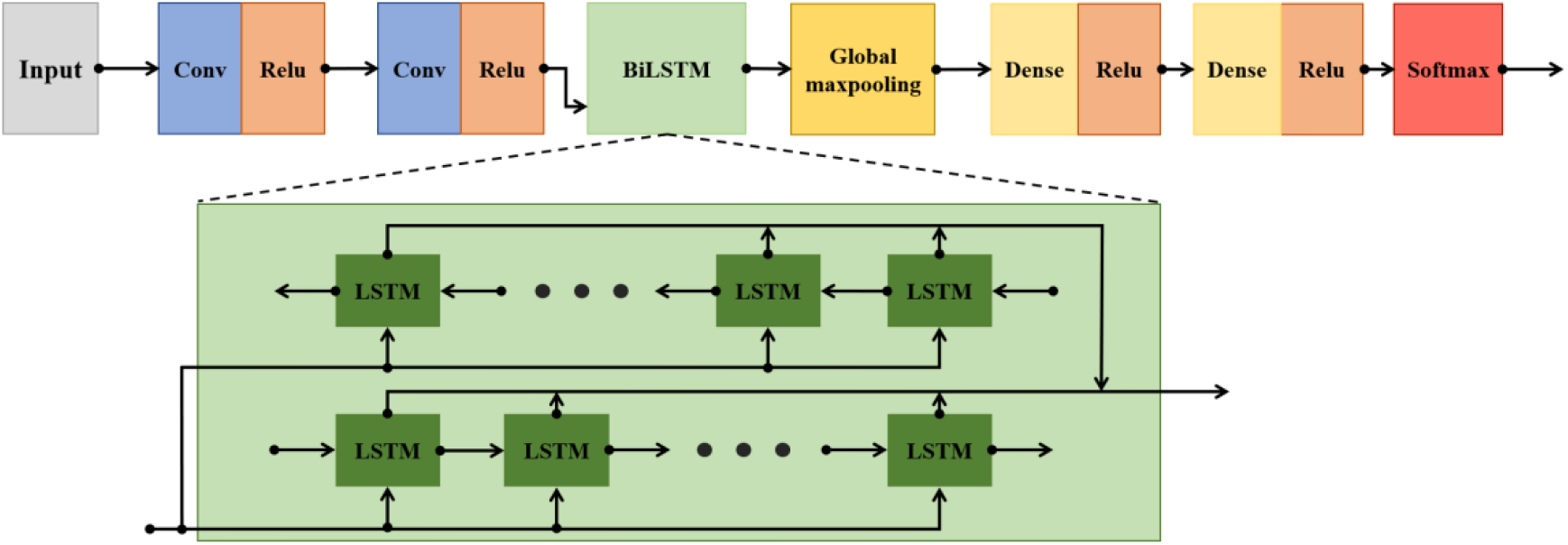
Architecture of the convolutional, long short-term memory, and fully connected deep neural networks.

**Figure. S2.**
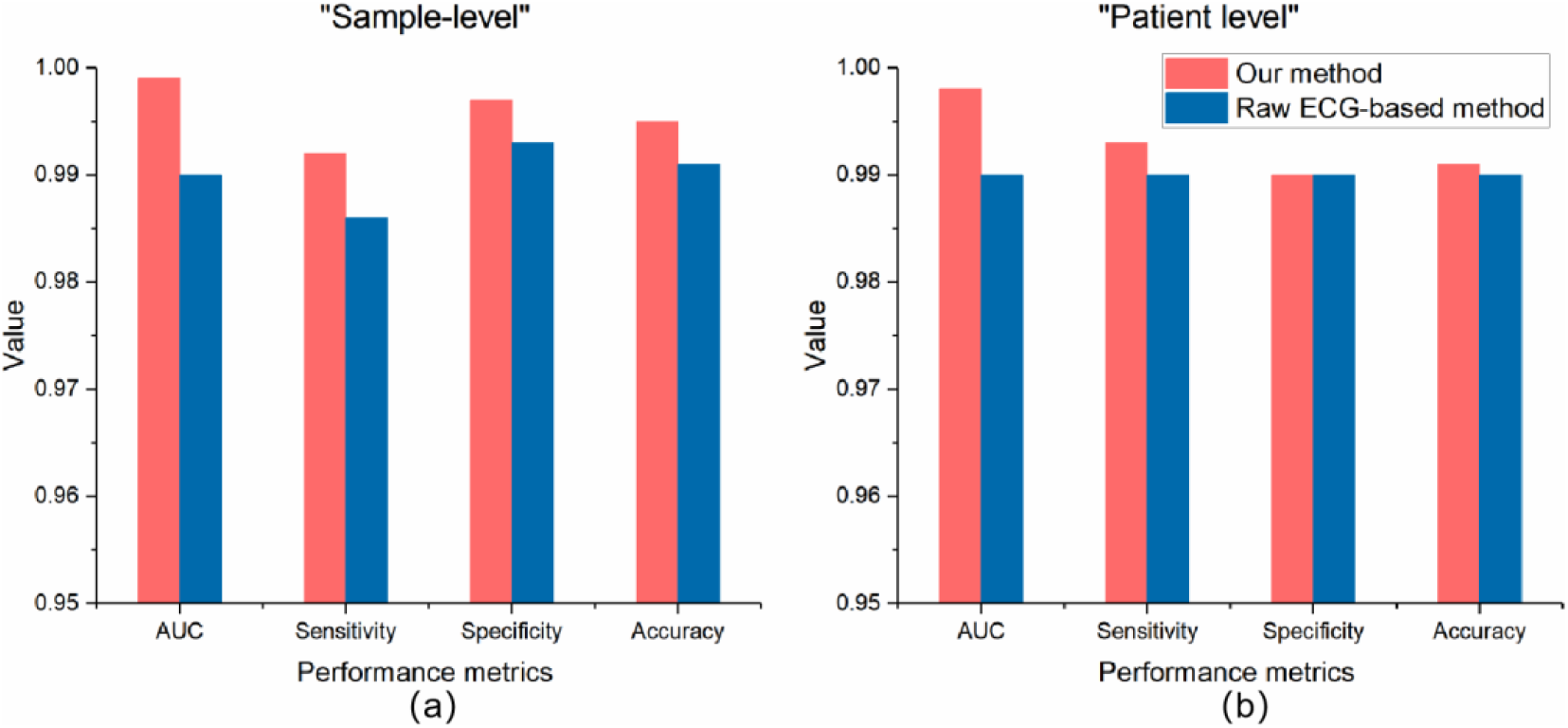
The comparison between our RR-interval-based method and the raw-ECG-based method in AF detection. (a) The results of AF detection at the “sample-level”. (b) The results of AF detection at the “patient level”. The red and blue bars represent the results of our RR interval-based method and the raw ECG-based method, respectively. AF = atrial fibrillation. AUC = area under the receiver operating characteristic curve.

**Figure. S3.**
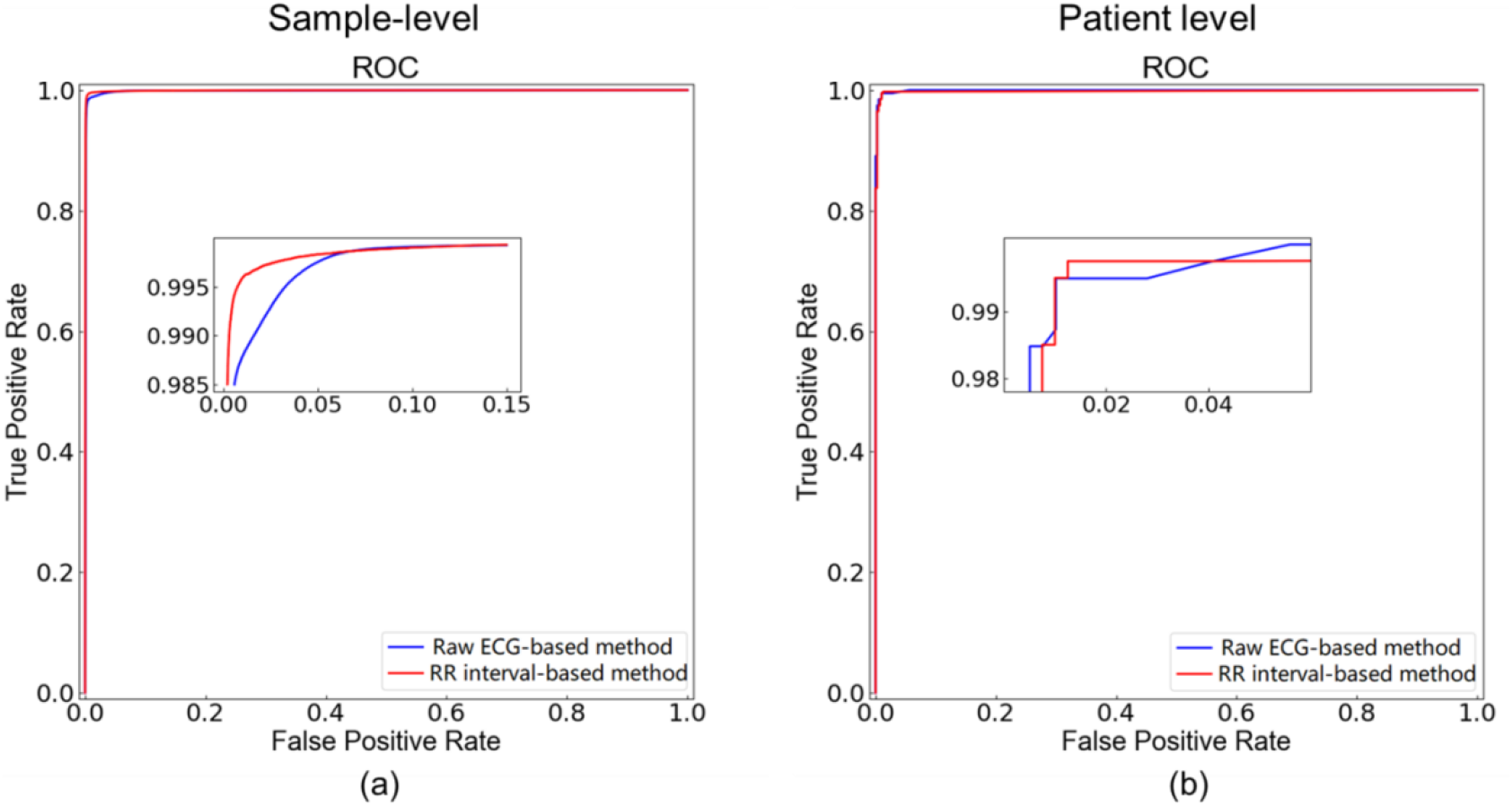
The ROC curves for the RR-interval-based and raw-ECG-based methods at the “sample-level” and “patient level” in the randomized clinical cohort. (a) The ROC curves for the “sample-level” analyses. (b) The ROC curves for the “patient level” analyses. The red and blue lines represent the RR interval-based and raw ECG-based methods, respectively. ROC = receiver operating characteristic.

**Figure. S4.**
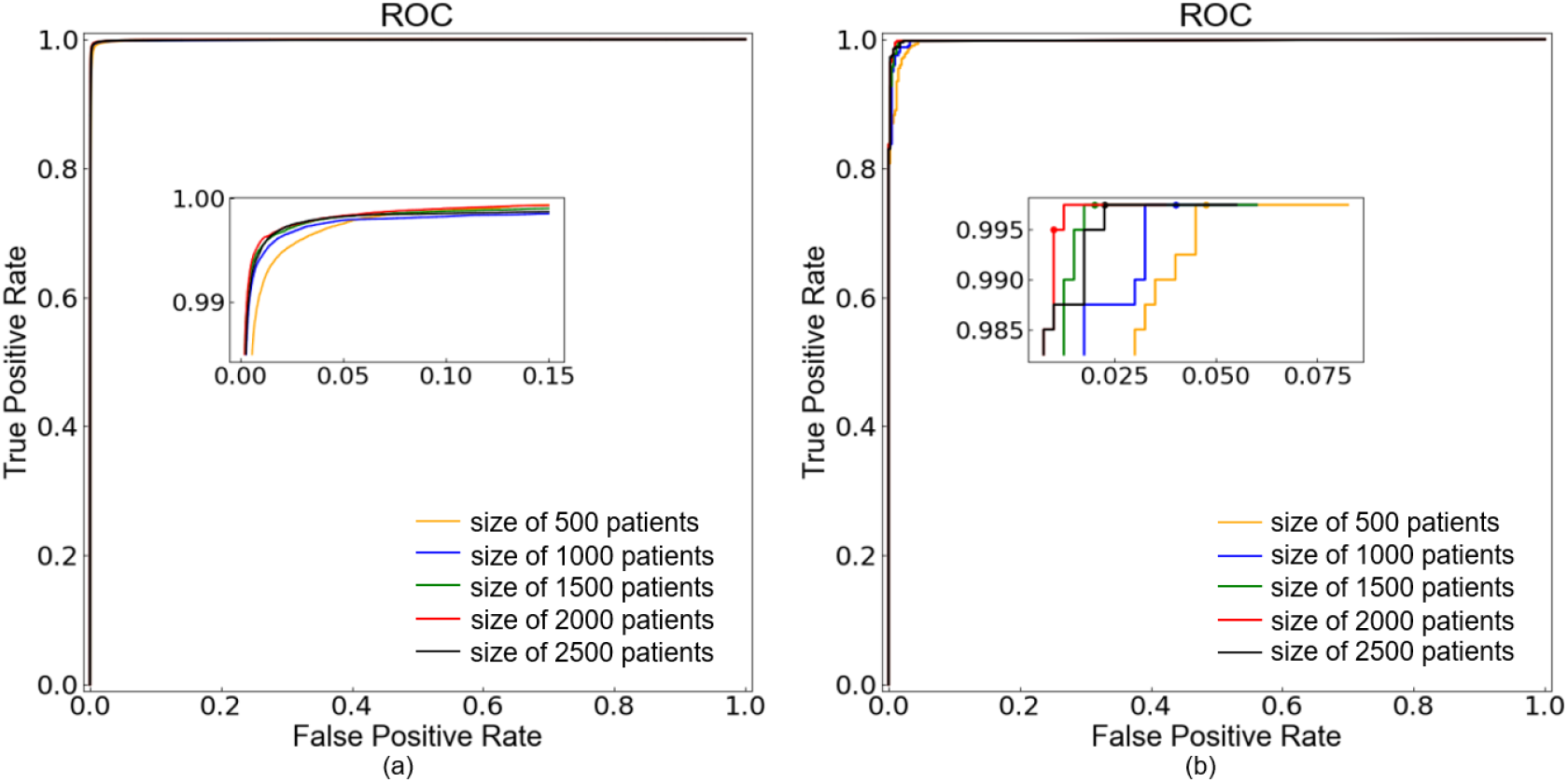
Effect of training dataset size on the detection performance. The yellow, blue, green, red and black lines represent the training dataset size of 500, 1000, 1500, 2000 and 2500 patients, respectively. (a) The receiver operating characteristic (ROC) curves for the “sample-level” analyses with different training dataset sizes. (b) The ROC curves for the “patient level” analyses with different training dataset sizes.

**Figure. S5.**
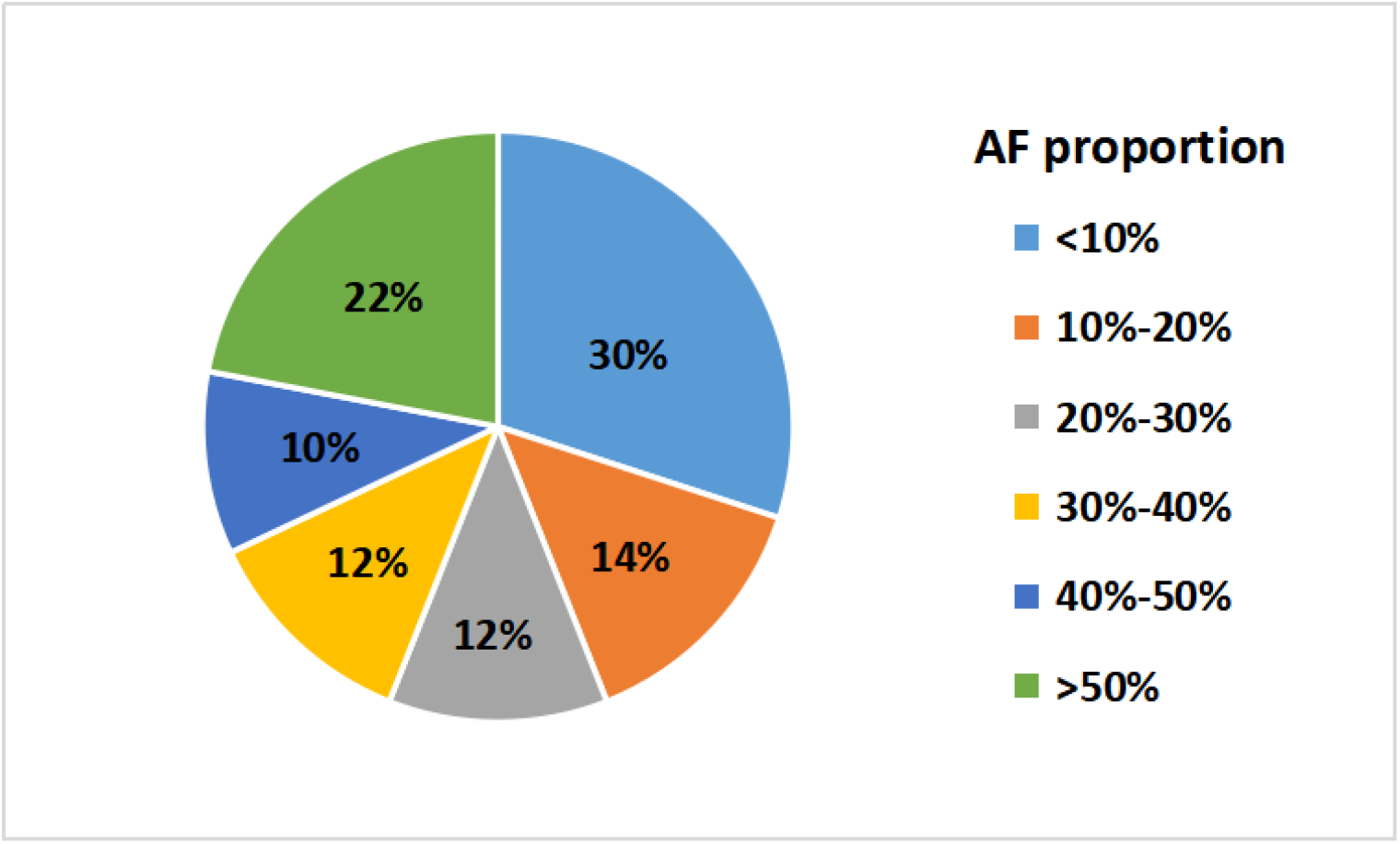
Distribution of 200 testing paroxysmal atrial fibrillation (AF) patients with different proportions of AF episodes in 24 h Holter recordings. The light blue, orange, gray, yellow, blue, and green represent the proportions of AF episodes of less than 10%, between 10-20%, 20-30%, 30-40%, 40-50%, and more than 50%, respectively.

